# Exploring MAPT-containing H1 and H2 haplotypes in Parkinson’s disease across diverse populations

**DOI:** 10.1101/2025.10.17.25337033

**Authors:** Paula Reyes-Pérez, Jia Wei Hor, Tzi Shin Toh, Arinola O Sanyaolu, Caroline B Pantazis, Thiago Peixoto Leal, Sheila Yeboah, Sara Bandres-Ciga, Huw R Morris, Mary B Makarious, Konstantin Senkevich, the Latin American Research Consortium on the Genetics of Parkinson’s Disease (LARGE-PD), the Global Parkinson’s Genetics Program (GP2), Hampton Leonard, Kajsa Atterling Brolin

## Abstract

Variation at the 17q21.31 locus, which contains the gene encoding microtubule-associated protein tau (*MAPT*), has been associated with neurodegenerative disorders, including Parkinson’s disease (PD). This highly complex locus is characterized by two broadly defined haplotypes: H1 and the inverted H2 haplotype. While H1 has been associated with an increased PD risk and is present in all ancestry populations, H2 is enriched in individuals of European ancestry. So far, few studies have explored the H1 association with PD in non-European ancestries. Here, we investigated the haplotype and subhaplotype frequencies of H1 and H2 in 20,507 PD patients and 11,841 controls across eleven different ancestry groups from the Global Parkinson’s Genetics Program (GP2) and the Latin American Research consortium on the GEnetics of Parkinson’s Disease (LARGE-PD). Our results strongly support the involvement of the H1 haplotype in PD risk in individuals of European ancestry, with additional evidence suggesting an association across diverse ancestry groups. Additionally, we observed significant variation in the H1 subhaplotype frequencies within populations, highlighting the complexity of this genomic region and the relevance of its study in diverse ancestries to gain a more comprehensive understanding of the role this locus plays in neurodegenerative disease risk.

## Introduction

Parkinson’s disease (PD) is an age-related progressive neurodegenerative disorder marked by clinical heterogeneity and driven by a complex interplay of genetic and environmental factors that vary across regions and contexts, influencing both its development and progression ^1,2^. Advances in the genetics of PD, particularly through large genome-wide association studies (GWAS) and meta-analyses, have identified numerous risk loci and candidate genes ^3–5^. Although many of these studies have predominantly focused on populations of European ancestry, efforts are increasing to include underrepresented non-European populations ^6–9^. In 2009, Simón-Sánchez and colleagues conducted a European GWAS in PD, including 5,074 patients and 8,551 controls, and identified two association signals linked to PD risk: one in *SNCA* and the other at the 17q21.31 locus ^10^. The same year, Satake and colleagues conducted a PD GWAS in individuals of Asian ancestry, including 2,011 patients and 18,381 controls, and did not find the association with variability at the 17q21.31 locus ^11^. This association also failed to be detected in the more recent and larger GWAS of South East Asian ancestry participants by Foo et al.^12^. However, the 17q21.31 locus has been consistently replicated in subsequent GWAS performed in European ancestry participants ^5^.

*MAPT*, located on chromosome 17q21.31, encodes the microtubule-associated protein tau, consists of 15 exons, and is primarily expressed in neurons, where it regulates axonal transport ^13–15^. The genomic architecture of the 17q21.31 region is highly complex, featuring a ∼1.8 Mb block of linkage disequilibrium (LD) defined by a 900 kb inversion and characterized by two main haplotypes, H1 and H2 ^13,14,16^. This locus represents the largest area of LD known in the human genome, and the inversion spans 15 genes as well as several non-coding RNAs and pseudogenes ^17,18^. The H1 haplotype exhibits normal genetic variation patterns, leading to multiple subhaplotypes ^14^. Single nucleotide variants (SNVs) can distinguish H1 from H2, with H1-specific SNVs generating several common H1 subhaplotypes ^16^. Numerous SNVs can be used to define the H1 vs H2 haplotypes, and H1 sub-haplotypes, but the most commonly used are rs8070723 and rs1052553, while sub-haplotype definitions are often based on rs1467967, rs242557, rs3785883, rs2471738, and rs7521, all located in *MAPT* ^18^.

The H1 haplotype is observed in all populations whereas the H2 haplotype (inverse orientation) is enriched in individuals of European ancestry, with a prevalence of about 20%. It is less common in African ancestry populations and nearly absent in East Asian ancestry populations, and shows very limited genetic variability ^14,17–19^. The H1 haplotypes have been associated? with an increased risk for several synucleinopathies, including PD, dementia with Lewy bodies, and multiple system atrophy ^14,16,20–22^. In contrast, the H2 haplotype is thought to reduce the risk for these neurodegenerative diseases ^4,14,16,23^. The H1 haplotype and H1c sub-haplotype are also strongly associated with the primary tauopathy Progressive Supranuclear Palsy (PSP), which involves the deposition of four-repeat predominant tau protein ^24^. There is some evidence that the H1 haplotype and H1c sub-haplotype increase the expression of *MAPT* and the inclusion of exon 10, providing a link between the genetics of PSP and neuropathology ^25,26^. However, many PD patients do not have tau pathology at post-mortem and it is uncertain whether *MAPT* itself explains the association between PD and the H1 locus. In fact, the haplotypes and variants at the locus have been shown to regulate gene expression of other genes at the locus, i.e., *KANSL1*, *CRHR1* and *LRRC37A/4*, in addition to *MAPT* ^27,28^. Although the 17q21.3 locus risk association is a prominent finding in PD genetics, the H1/H2 association has been largely reported in European populations, likely related to the frequency and sample size of the non-European studies and the low H2 background allele frequency in Africa and Asia ^17,29^. Given the limited research on 17q21.3 haplotypes in PD among non-European cohorts, this study aims to investigate the frequency and associations of the H1 and H2 haplotypes with PD across diverse ancestries using data from the Global Parkinson’s Genetic Program (GP2, https://gp2.org/) ^30^ in 20,507 PD patients and 11,841 controls and the Latin American Research consortium on the Genetics of Parkinson’s Disease (LARGE-PD) ^8^, consisting of 775 PD patients and 673 controls. Association analysis in non-European populations may help define the causal alleles and by examining these haplotypes across a broad range of ancestry groups, we seek to enhance our understanding of their role in PD and provide new insights into the genetic underpinnings across diverse populations.

## Methods

### Samples

We investigated the frequency of the H1 and H2 haplotypes using genotyping data from the GP2 release 7 (DOI: https://zenodo.org/records/10962119) ^31^, consisting of 20,507 PD patients and 11,841 controls from ten different ancestry groups: African American (AAC), African (AFR), Ashkenazi Jews (AJ), Admixed American/Latin American (AMR), Central Asian (CAS), Complex Admixture History (CAH), East Asian (EAS), European (EUR), Middle Eastern (MDE) and South Asian (SAS). Additionally, we investigated the haplotype frequencies in LARGE-PD ^32^. The LARGE-PD cohort includes 775 Parkinson’s disease (PD) patients and 673 controls, recruited from nine clinical sites across South America, and represents a Latino or American admixed population comparable to the GP2-AMR cohort. Although the LARGE-PD cohort is can be classified under the AMR population label, for clarity and ease of reading, we will refer to GP2-AMR as “AMR” and LARGE-PD as “LARGE-PD” throughout this work. After filtering out related individuals and individuals with missing genotype data for the investigated H2-tagging SNV, a total of 19,921 PD patients and 11,401 controls remained for the analysis. The breakdown of the numbers of PD patients and controls is listed in Table 1, and a summary of demographics can be found in Supplementary Table 1.

**Table 1:**
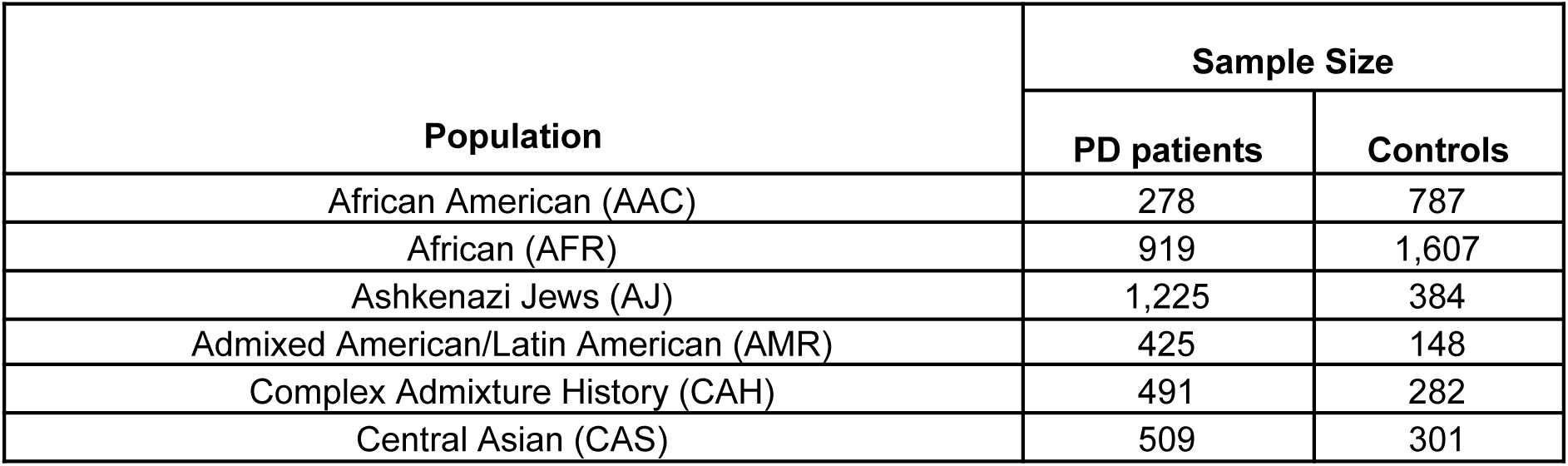
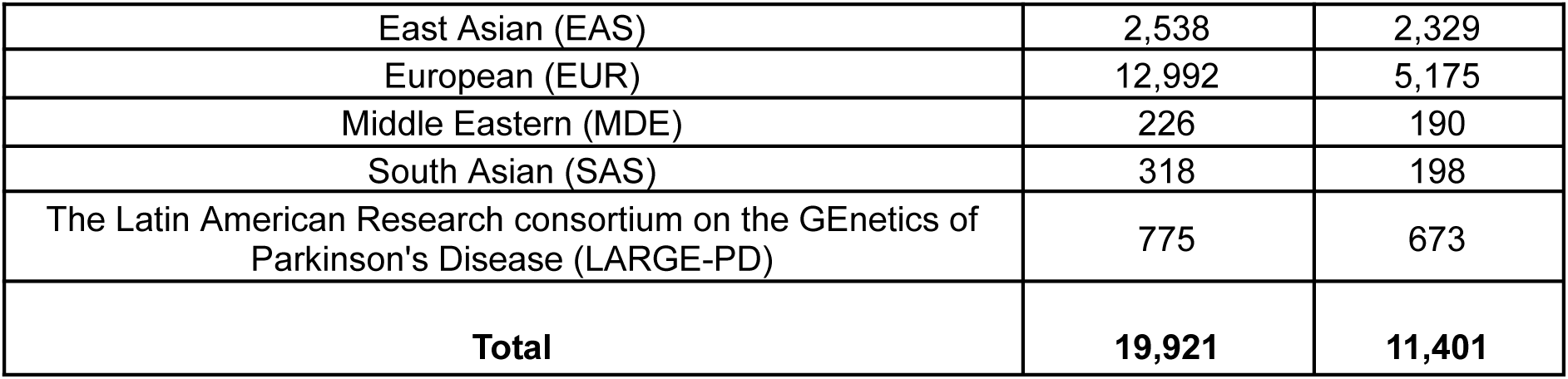
Number of PD patients and controls from each ancestry included in the study.

### Power calculation

Power calculations were done using the Genetic Association Study (GAS) Power Calculator (https://csg.sph.umich.edu/abecasis/cats/gas_power_calculator/) using an additive model, a significance level of p=0.05 and a disease prevalence of 0.5% with the available sample sizes and the observed minor allele frequency (MAF) in each ancestry group, ranging from 0.26% in EAS to 22.99% in AJ. Additionally, the estimated number of samples needed to reach 80% power at the effect size (odds ratio [OR]) observed in the EUR population was calculated for the observed MAF in the different ancestry groups (Supplementary Table 2).

### Genetic data and analyses

Genotyping (Illumina NeuroBooster Array [NBA]) ^33^, sample and variant quality control (QC), imputation, ancestry prediction, and processing in GP2 were previously performed using GenoTools v1.0, and have been described elsewhere (https://github.com/dvitale199/GenoTools) ^34^. All analyses using GP2 data were conducted on the cloud-native platform, Terra Community Workbench (https://app.terra.bio/), and were performed separately for all ancestry groups. The LARGE-PD data was genotyped using the Illumina Multi-Ethnic Genotyping Array ([MEGA], ^8^. Genotyping QC and relationship control were performed using the LARGE-PD QC pipeline (https://github.com/MataLabCCF/GWASQC). This pipeline was specifically developed to perform QC in admixed populations, minimizing unnecessary sample loss. . Imputation for both GP2 and LARGE-PD was done using the TOPMed reference panel as described ^35^.

17q21.3 H1/H2 haplotypes were investigated using the H2-tagging SNV rs1052553 (chr17:45996523:A:G) from the raw genotyping data according to the following allele combinations: AA representing the H1/H1 haplotype, AG the H1/H2 haplotype, and GG the H2/H2 haplotype. Allele and haplotype frequencies, Hardy Weinberg equilibrium (HWE) and ancestry specific associations with PD risk and PD age at onset (AAO) were analyzed using PLINK v1.9 and v2.0 with and without adjusting for (i) sex and PC1-5 and (ii) sex, age, and PC1-5 (PD risk only). Multiple-testing correction was applied to the p-value threshold for the number of tests, resulting in Bonferroni-corrected p-value of 0.05/11=0.0045. HWE was calculated in the control group for all ancestry populations. Plots for the frequencies across ancestries were done using *ggplot* in R version 4.1.2, while association results were visualized using the *forestplot* package in Python version 3.10.9. Potential differences for AAO between H1 and H2 carriers were evaluated using the Mann–Whitney U test as the data was not normally distributed, and a linear regression adjusted for sex and PCs 1-5 was performed.

Six tagging variants in the locus (chr17:45908813:G:A - rs1467967; chr17:45942346:G:A - rs242557; chr17:45977067:A:G - rs3785883; chr17:45998697:C:T - rs2471738; chr17:46003698:A:G - rs8070723; and chr17:46028029:A:G - rs7521) were used to construct the most common subhaplotypes as previously described using the Pittman SNV-based nomenclature ^36,37^. These variants have been reported to capture 95% of the common 17q21.3 haplotype diversity in Europeans ^37^, where five of these variants are H1-specific. To further distinguish the H2 subhaplotypes (H2 and H2D), the SNVs rs1052553 (chr17:45996523:A:G - rs1052553, chr17:46724418:G:A - rs199451, and chr17:46751565:G:A - rs199533) were used ^18,38^. We required that all three SNVs match the expected haplotype.

Since not all variants were captured during genotyping, we utilized multi-ancestry imputed genotyping data to investigate the H1 subhaplotypes. As genetic imputation uses known haplotypes in a reference population, we investigated the subhaplotypes in a subset of individuals where both whole genome sequencing (WGS) data (GP2, release 6, ^39^) and imputed genotype data were available in GP2 to evaluate the accuracy. This was evaluated in 150 individuals (99 PD patients and 51 controls) of the AFR ancestry group and 533 individuals (452 PD patients and 92 controls) of the EUR ancestry group.

To further elucidate the relationship between 17q21.3 subhaplotypes and PD, we employed GenoML ^40^, an automated machine learning tool for genomics in a per-ancestry analysis. The imputed pVCF was subsetted to the six tagging SNVs and then recoded to create a PLINK .raw file. Each SNV was coded as follows: 0, for being homozygous for the reference allele, 1 for heterozygous, and 2 for homozygous for the alternate allele. Samples that were heterozygous at tagging SNVs were coded to the alternate allele. After assigning a subhaplotype to each sample, the resulting data were converted into a binary representation using one-hot encoding to prepare for input into GenoML. To rank the relative importance of a subhaplotype in predicting PD per ancestry, we used GenoML’s feature selection flag, which calls on Sci-Kit Learn’s Extra Trees Classifier ^41^. In this model, the data is split at decision tree nodes based on the presence or absence of the subhaplotype used to make the split. The classification error is the Gini impurity, which quantifies the likelihood of incorrectly classifying a randomly chosen subject from the dataset if it were labeled according to the class distribution within a node. A lower Gini impurity indicates that the node predominantly contains individuals from a single class. The importance of a subhaplotype is computed as the normalized total reduction in classification error that results from including this subhaplotype in the model— also known as the Gini importance. The higher the Gini importance, the more informative the subhaplotype is in distinguishing between PD and controls.

Additionally, to get a general view of the potential association between the 17q21.31 genomic region and PD, we extracted the region (chr17:42,800,001-46,800,000 [UCSC Genome Browser, GRCh38/hg38]) and conducted association and logistic regression analyses using PLINK v2.0, adjusting for sex, age, and the first 5 PCs, filtering by MAF>1%. A regional plot spanning the 17q21.31 locus was visualized employing the *topr* tool version 1.1.8 ^42^ in R v4.4.0.

## Results

The rs1052553-AA genotype, representing the H1/H1 haplotype, was the most frequent across all ancestry populations, followed by H1/H2 (AG) and H2/H2 (GG) (Figure 1, Supplementary Table 3). The H2 haplotype frequency varied across the ancestry groups, ranging from 0.1% in PD patients and 0.4% in controls in the EAS ancestry population to the highest frequencies in the AJ population with 21.0% among PD patients and 25.0% in controls. In most groups, H2 was more common in controls than in PD patients, except in the AMR and CAH ancestry groups, where H2 was more common among PD patients, MAF_H2_=14.2% vs 14.5% in AMR and MAF_H2_=10.5% vs 12.6% in CAH. The rs1052553 variant was in HWE in the control group in all populations except for the MDE population (p=0.004) (Supplementary Table 4).

**Figure 1:**
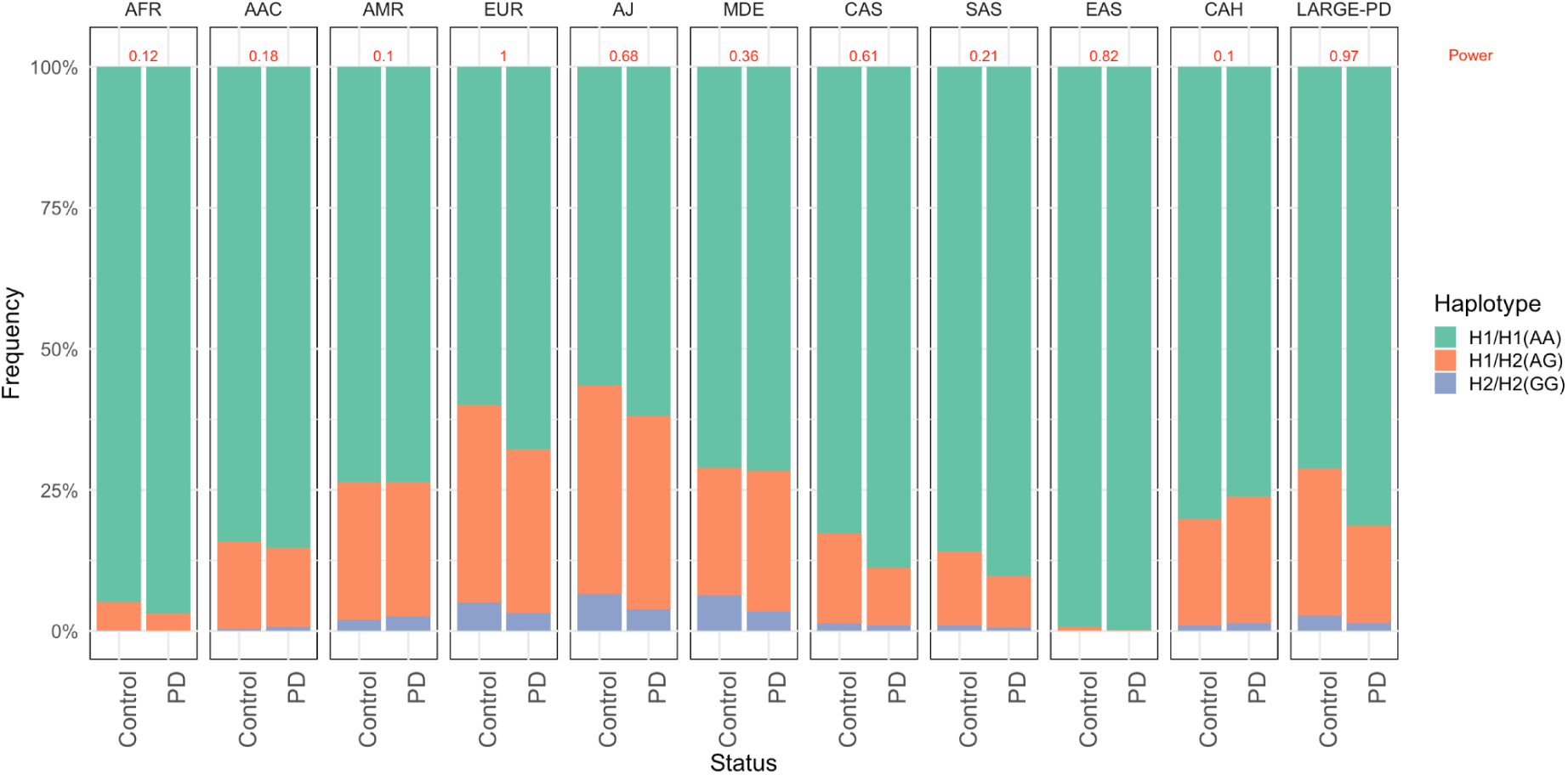
H1/H2 allele/haplotype frequency (rs1052553) in PD patients and controls in African (AFR), African American (AAC), Admixed American/Latin American (AMR), European (EUR), Ashkenazi Jews (AJ), Middle Eastern (MDE), Central Asian (CAS), South Asian (SAS), East Asian (EAS), Complex Admixture History (CAH) populations and the Latin American Research consortium on the GEnetics of Parkinson’s Disease (LARGE-PD). In red at the top of each plot, the power for each ancestry is shown.

A nominal statistically significant difference for the allele/haplotype frequency between PD patients and controls was observed in the five ancestry populations AFR (OR=1.76, 95% confidence interval [CI]=1.14-2.70, p=0.0095), AJ (OR=1.256, 95% CI=1.04-1.52, p=0.012), CAS (OR=1.58, 95%CI=1.07-2.30, p=0.016), EAS (OR=3.28, 95%CI=1.30-8.27. p=0.0077), and EUR (OR=1.35, 95%CI=1.28-1.43, p=2.29E-26), as well as in the LARGE-PD cohort (OR=1.67, 95%CI=1.34-2.08, p=4.6E-6) (Supplementary Table 5, Supplementary figure 1).

Additionally, after adjusting the regression for age, sex, and PC1-5, a statistically significant association between the H1 haplotype and PD was only observed in the EUR population (OR=1.33, 95%CI=1.24-1.43, p=2.4E-15) and the LARGE-PD cohort (OR=1.51, 95%CI=1.19-1.93, p=0.000745) (Supplementary Table 6, Supplementary Figure 2). Complete-case analyses were used for the multivariate logistic regression, resulting in a substantial loss of participants in several ancestry groups due to missing age data. To increase the sample size and hence the statistical power, we additionally ran a multivariate logistic regression only including sex and PC1-5 as covariates which generated nominal significant associations in the AJ (OR=1.27, 95%CI=1.05-1.54, p=0.0149) and CAS ancestries (OR=1.57, 95%CI=1.08-2.26, p=0.0169), in addition to the significant associations in the EUR population (OR=1.32, 95%CI=1.25-1.30, p=1.31E-22), and the LARGE-PD cohort (OR=1.51, 95%CI=1.19-1.91, p=0.00067) (Supplementary Table 7, Figure 2).

**Figure 2.**
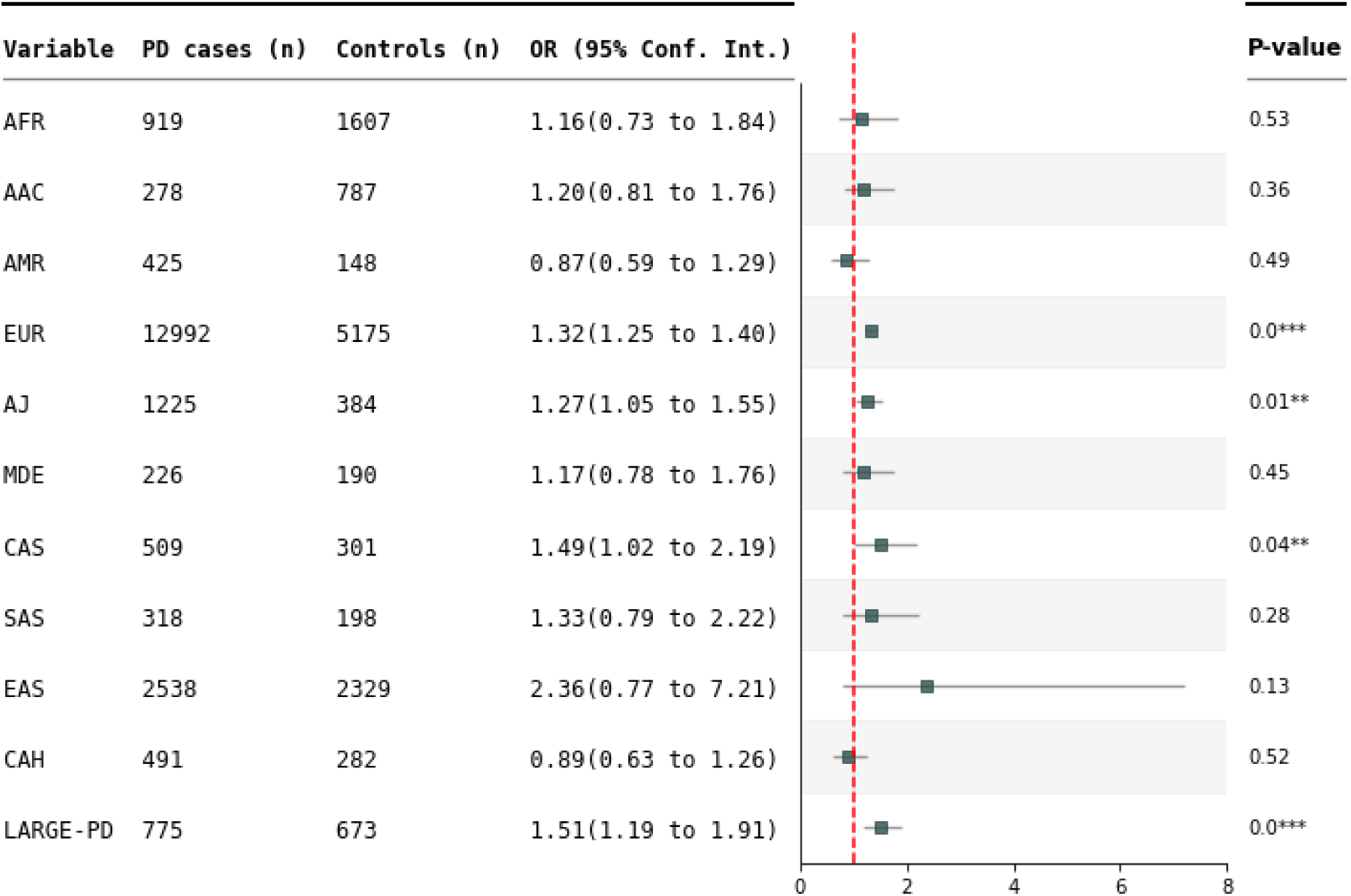
Forest plot showing odds ratio (OR) estimates with a 95% confidence interval for the *MAPT* containing H1 haplotype with Parkinson’s disease in the GP2 and LARGE-PD cohorts. N shows the sample size per population after adjusted regression (sex and PC1-5). P-values are marked with stars, with * indicating p < 0.05, ** corresponding to p < 0.01 and *** show p < 0.001). A dashed red line indicates OR = 1.

No differences were observed for PD AAO between H1 and H2 haplotype carriers in any ancestry group (Supplementary Table 8). A nominal association (not passing Bonferroni significance) between the H1 haplotype and AAO was observed in a linear regression adjusted for sex and 5PCs in the AJ and LARGE-PD groups (Supplementary Table 9), with the H1 haplotype being associated with a ∼1.3 and ∼3.2 years earlier AAO compared to the H2 haplotype (AJ: beta = -1.368, SE = 0.657, p = 0.0374, LARGE-PD: beta = -3.175, SE = 1.253, p = 0.0115).

The subhaplotype analyses included six tagging variants, which capture the H2 haplotype along with the variation patterns of the H1 subhaplotypes. The variants were in HWE in all populations with a few exceptions (chr17:45908813:G:A in AFR, chr17:45942346:G:A in AJ, chr17:46003698:A:G in AAC and MDE, and chr17:46028029:A:G in LARGE-PD) (Supplementary Table 10). To investigate the accuracy of the genomic imputation for the data used in the subhaplotype analysis, we evaluated the subhaplotype pattern in a subset of individuals of the AFR and of the EUR ancestry group for which both WGS and imputed genotyping data was available. We observed a similar pattern for both methods, with minor allele frequencies discrepancies (<0.4 percentage points difference in EUR and <2.1 percentage points in AFR [Supplementary Figure 3 and 4]).

A large variation in the H1 subhaplotype frequencies was observed between several ancestry populations (Figure 3, Supplementary Table 11). EUR, AJ, and MDE populations showed a similar pattern, with H1b subhaplotype being the most frequent, followed by H1c. In contrast, these subhaplotypes were rare in the EAS population (1.6% and 0.4%, respectively), where H1 subhaplotype M was the most common, similar to CAS population. The highest frequency of the H1J subhaplotype was seen in AFR (17.1%) and AAC populations (13.8%) followed by EAS (10.7%) with the lowest in EUR population (1.1%). A statistically significant difference in the frequency between PD patients and controls was found only for H2a subhaplotype in EUR (p=5.01E-24) and the four H1 subhaplotypes H1b (p=0.0003), I (p=0.0009), L (p=1.6E-05), and M (p=4.2E-07) in EAS after Bonferroni correction (p=0.05/33=0.0015).

**Figure 3:**
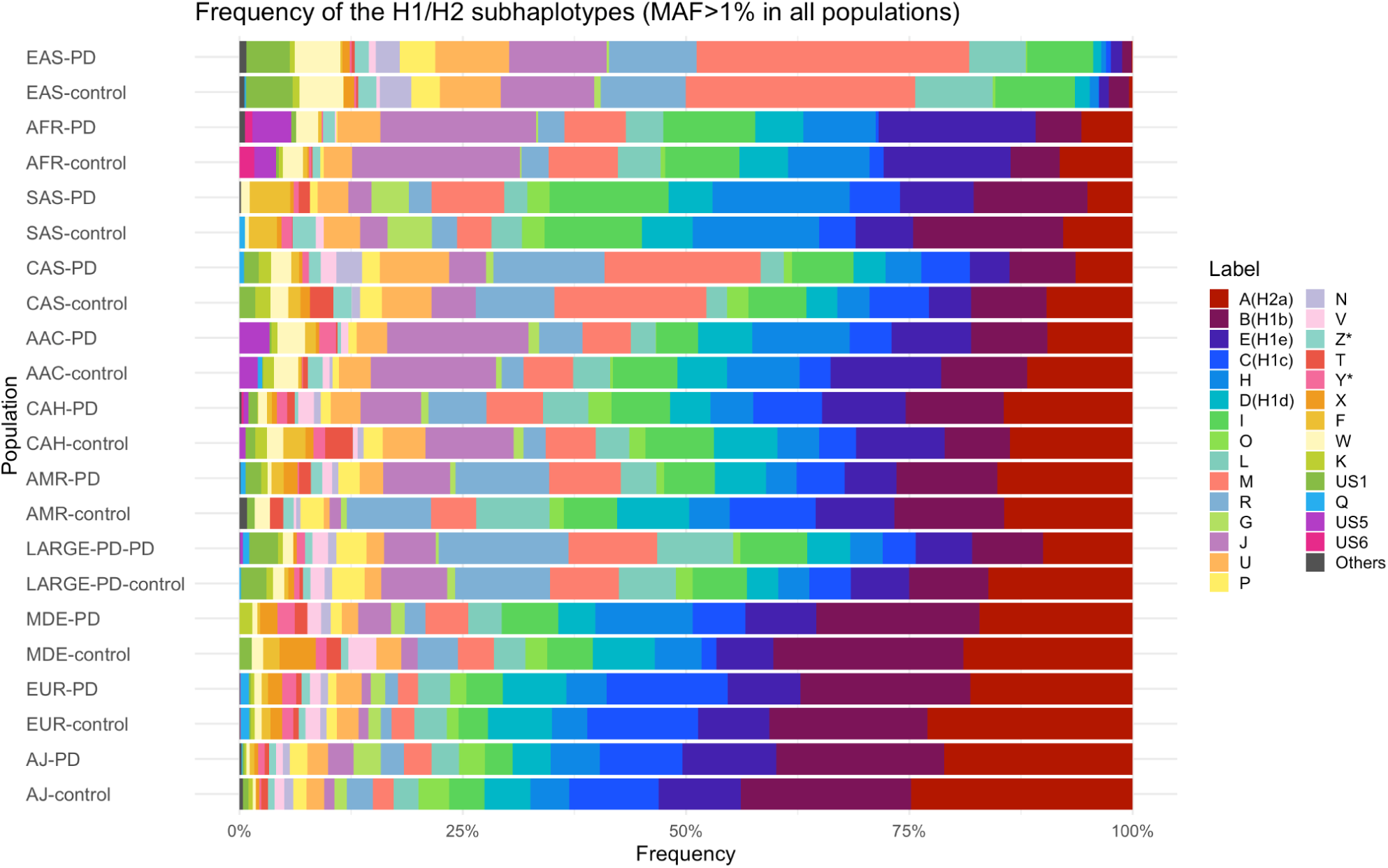
Frequency of H1 and H2 subhaplotypes in PD patients and controls. Showing the frequency (MAF>1%) of the H1 and H2 subhaplotypes in the ancestry groups African (AFR), African American (AAC), Admixed American/Latin American (AMR), European (EUR), Ashkenazi Jews (AJ), Middle Eastern (MDE), Central Asian (CAS), South Asian (SAS), East Asian (EAS), and Complex Admixture History (CAH) populations. US: Unspecified subhaplotype. Plots are ordered by the frequency of A(H2a) subhaplotype in controls.

After ranking the subhaplotypes by Gini importance score, none of subhaplotypes with the highest importance scores overlapped between the ancestries (Supplementary Table 12). In the EUR ancestry group, the H2A subhaplotype was ranked at twice as important than the second and third most important subhaplotypes, the US6 and H1C subhaplotypes, further emphasizing that the H2a subhaplotype is most important in distinguishing between PD patients and controls in the EUR ancestry group. In the EAS ancestry group, the L subhaplotype ranked three times higher than the second-ranking haplotype (the Y* subhaplotype), indicating the importance of the L subhaplotype in this ancestry group.

For subhaplotypes in H2, the H2a (SNV-based nomenclature, ^37^) was seen to be the most common in all ancestry groups. The same SNV tagging the H2 (H2a) haplotypes in the analysis (rs8070723) has also been described to tag the H2D subhaplotype (CNV-based nomenclature, ^18^). Further analyses of the H2 haplotype showed that the H2D haplotype was most common in all ancestry groups with the exception of the AFR population where the H2.1 subhaplotype was more frequent (Table 2).

**Table 2:**
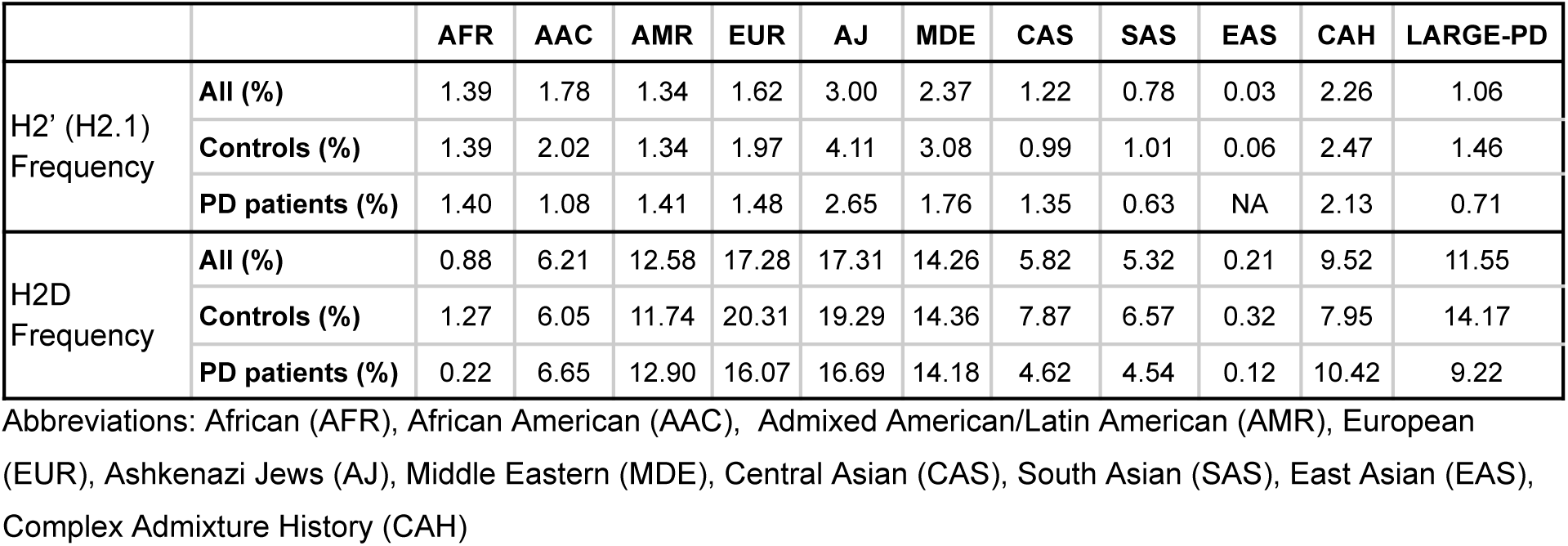
Frequency of the major H2 subhaplotypes in the GP2 ancestry populations and LARGE-PD.

We additionally generated regional plots to explore the association signals in the 17q21.31 region across the different ancestry groups. In the EUR population, which had a considerably larger sample size compared to other groups, we observed significant associations between multiple variants and PD throughout the region of *MAPT*. The pattern of significant variants is indicative of the strong LD in this region and a similar LD block structure was observable in the plots for the other ancestry populations; however, no significant association was observed between PD and any variant in the region at genome-wide significance level (Figure 4, Supplementary Figure 4).

**Figure 4.**
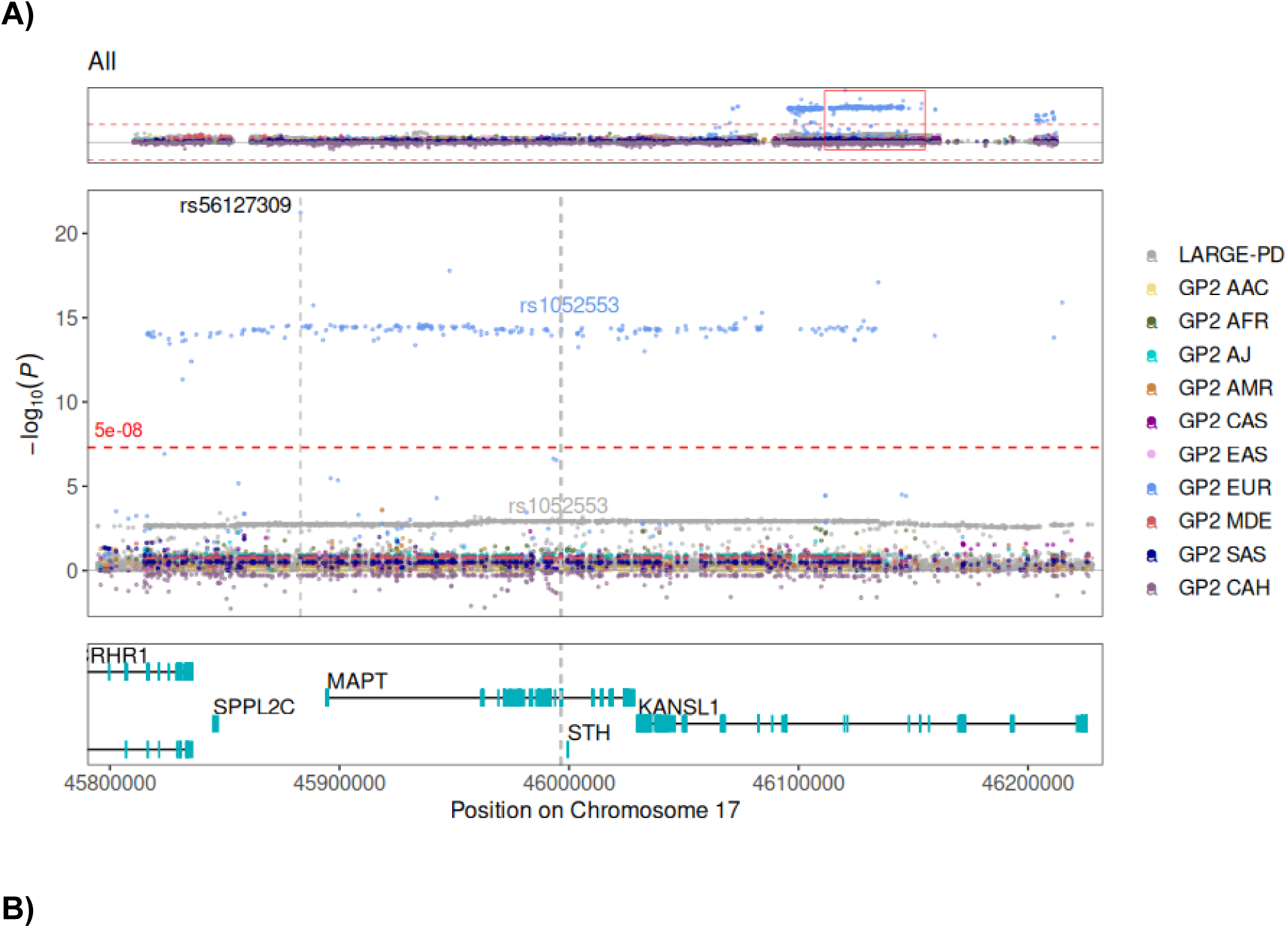
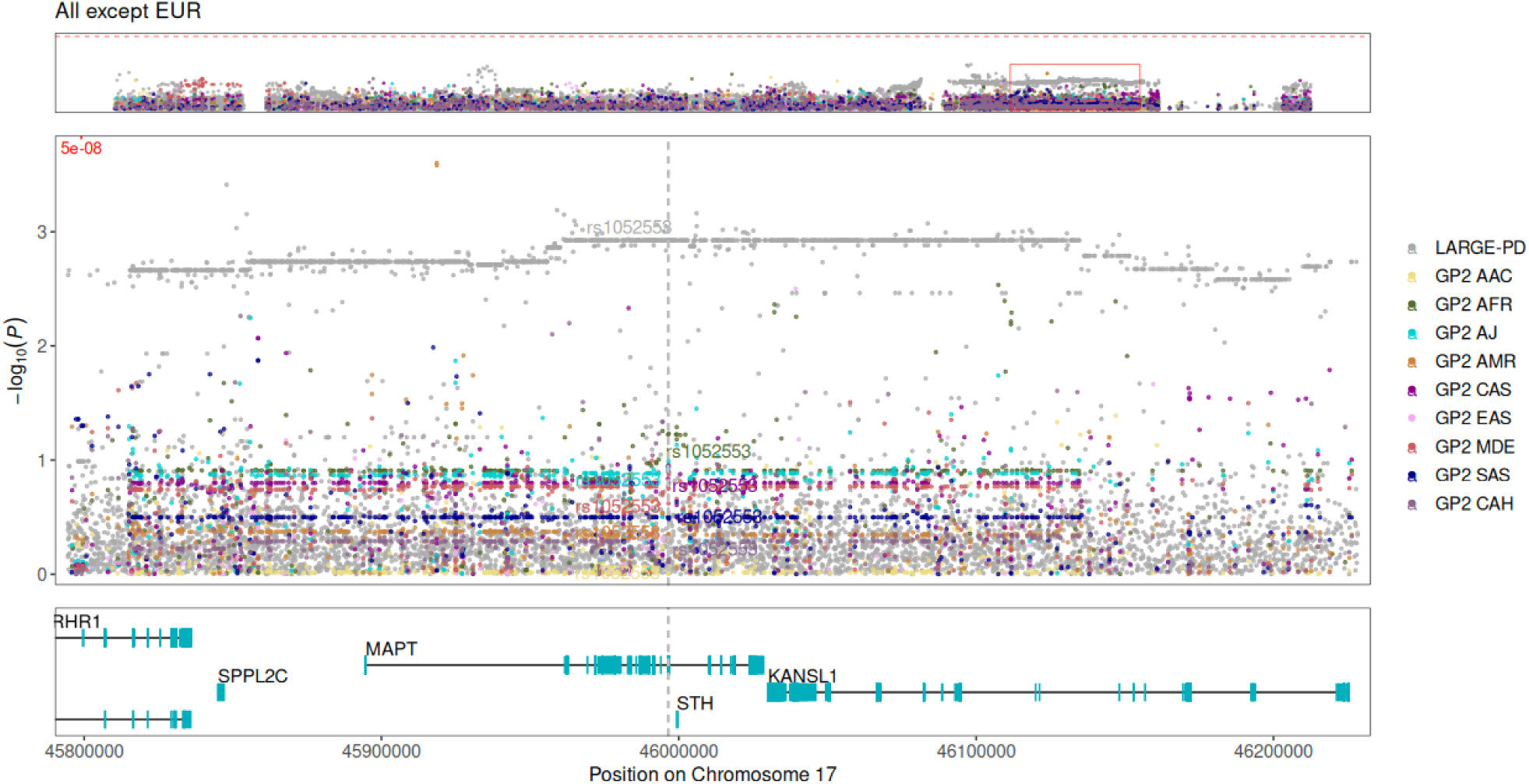
Regional plot of variant associations at the *MAPT* locus. (A) Variant association results across all ancestry groups (AAC, AFR, AJ, AMR, CAS, EAS, EUR, MDE, SAS and CAH) from the GP2 and LARGE-PD cohorts. (B) The same analysis in which the EUR population was removed to improve visualization of associations in the remaining populations. Association results are derived from an adjusted regression model with MAF filter of >1%. The y-axis represents –log₁₀(p) values of variant associations, while the x-axis displays genomic positions. In each plot, the upper panel presents the full 17q21.31 genomic region (chr17:42,800,001–46,800,000, UCSC Genome Browser, GRCh38/hg38). The middle panel zooms in on the *MAPT* region (chr17:45,794,527–46,228,334, with significant variants annotated by their rs IDs. The H2-tagging SNV (rs1052553) is marked by a vertical gray dotted line. The lower panel displays protein-coding genes adjacent to *MAPT*. The horizontal red dashed line represents the genome-wide significance threshold.

## Discussion

This study provides a comprehensive investigation of the *MAPT*-containing H1 and H2 haplotypes in PD across multiple ancestry groups. Our findings highlight the population-specific variability in haplotype frequencies and their association with PD risk, underscoring the importance of studying diverse cohorts in PD genetics. We observed that the H1 haplotype was present in all ancestry groups, while the H2 haplotype was significantly more frequent in EUR and AJ populations and nearly absent in the EAS ancestry population. This finding aligns with prior reports describing the H2 haplotype as a predominantly European marker with limited genetic variability ^18,43,44^.

Earlier studies of the H1 haplotype and PD have yielded mixed results. Several European cohorts reported an association of the H1 haplotype with PD, including Greek (OR:1.6, 95% CI: 1.1–2.2;) ^45^, Serbian (OR:2.0, 95% CI: 1.3–3.2) ^46^, Norwegian (OR:1.9, 95% CI: 1.3–2.6)^47^ and U.S. samples of European descent (OR:1.5, 95%CI: 1.3–1.7) ^5^, yet no signal was confirmed in a German cohort (36). Additionally, GWAS in individuals of European ^5,10^ and Latin American ancestries ^8^ have identified variants in the 17q21.31 region as associated with PD. Also a recent PD GWAS in the Indian population observed a borderline genome-wide significant signal at the *MAPT* locus with the H1/H2 haplotype tagging variant rs8070723 and identified a H2 haplotype frequency of 4% in the Indian population ^48^. In contrast, no association was observed in a Mexican mestizo population ^49^, in the Nigeria Parkinson’s Disease Research network cohort ^29^ or in Asian population GWAS ^11,12^. Due to the complex LD structure, the *MAPT* locus was excluded in the multi-ancestry PD GWAS by Kim *et al* ^9^ and the high LD makes it difficult to identify potential specific causal variants in the region ^18^. Haplotype-based analyses are therefore essential to provide a more comprehensive understanding of the genetic variability in the region.

We show that the H1 haplotype is associated with PD in multiple ancestry groups with an effect size ranging from OR=1.27 to OR=1.51, with the significance of the association varying in relation to the H2 allele frequency. We anticipate that the previous report of a lack of association is due to the sample sizes and that, as the sample size from diverse ancestry groups grows in GP2, the H1 haplotype will be associated with PD in all populations reflecting a common pathogenic pathway to PD. Supporting this is the observation of the H1 haplotype being more common among PD patients than controls in all ancestry groups with the exception of the GP2-AMR and the CAH ancestries (with a similar MAF for PD patients and controls). The conclusions that can be drawn from the CAH ancestry group are restricted by the limited sample size (491 PD patients and 282 controls) and the CAH ancestry group consists of highly admixed individuals from various backgrounds which would require special consideration and analysis to disentangle the genetic architecture of this locus. Due to varying MAF in the different ancestry groups, sufficient power to detect a true association was only estimated for EUR, EAS and LARGE-PD. To forecast the number of PD patients and controls needed to achieve 80% power in the different ancestry groups we performed power calculations assuming a similar effect size as the EUR ancestry group (OR=1.32). As an example, in the AFR ancestry group, where the frequency of the H2 haplotype is ∼2.1%, we estimate that we would need ∼4,360 PD patients and the same number of controls to reach significance at p=0.05.

We show that there is much greater haplotypic diversity across the 17q21.31 locus in non-European population groups, enabling fine mapping of functional alleles in different populations. While the H2 haplotype is most common in European populations, different H1 subhaplotypes, notably H1J and H1M, are more common in AFR and EAS, respectively. A potential association was only observed for four subhaplotypes in the EAS ancestry group, with the L subhaplotype ranking as the most important feature whereas the H2a subhaplotype had the highest rank in the EUR ancestry group. This could give us an indication of the subhaplotypes of highest importance in PD, but further analyses are needed to fully elucidate this. We additionally show that the H2D haplotype was most common in all ancestry groups with the exception of the AFR population where the H2’ (H2.1) subhaplotype was more frequent (although at low MAF), which is likely an ancestral H2 haplotype and has previously been reported to be enriched in African hunter-gatherer groups as compared to West Africans populations ^38^.

The four major sub-haplotypes of H1 and H2 have been described to be H1’, H1D, H2’ and H2D based on the inversion and copy number status of *KANSL1* ^38^. H1’ and H2’ carry no duplications (also defined as H2.1) whereas H1D spans a 205-kb duplication and H2D a shorter (155-kb) duplication. As CNVs were not investigated here and no common SNVs have been identified that could distinguish H1′ from H1D haplotypes ^38^, we relied on the SNV-based nomenclature from Pittman et al. to distinguish H1 subhaplotypes ^37^. However, for H2, this method only describes the H2a subhaplotype, and the same tag SNVs also tag the H2D subhaplotype ^18^. In contrast to H1D, the H2D can be defined based on multiple SNVs in the region while other SNVs located in *KANSL1* describe the H2’ subhaplotype. Pedicone and colleagues have suggested a hybrid nomenclature for subhaplotypes using Steinberg CNV-based nomenclature merged with Pittman SNV-based nomenclature separated by underscore ^18^. As the authors note, CNV- and SNV-based nomenclatures do not completely overlap and should be considered independently. Due to the absence of CNV data for H1, we relied solely on SNV-based methods.

Our study has several limitations. Firstly, although the study encompasses a substantial and diverse dataset, some demographic groups are still underrepresented, which may affect the applicability of the findings across all populations. Additionally, using other methods in the future such as SHAPEIT5 would be of interest to estimate subhaplotypes in a more unbiased approach, with better estimates of rare variants ^50^. We also observed missing data on key covariates, such as age, in a significant portion of individuals, which reduced the sample size and analytical robustness, resulting in that the results have been reported without adjusting for age. Moreover our subhaplotype analysis is based on imputed genotyping data. While this method is generally reliable, it can introduce some uncertainty in haplotype determination, especially in populations with less comprehensive linkage disequilibrium reference panels. To mitigate this issue, we compared results with WGS data, which revealed only minimal discrepancies. In addition, occasionally, the investigated variants deviated from HWE which is believed to be a consequence of the structural complexity of this region ^18,51^. Furthermore, analyses were conducted using datasets from GP2 and LARGE-PD cohorts, which employed different genotyping panels (NBA^33^. vs MEGA^8^), which may introduce inconsistencies in variant coverage and affect the comparability of results across cohorts. Finally, our study did not investigate CNVs, which are known to contribute to the genetic diversity of the 17q21.31 region and its association with PD ^38^.

In conclusion, our study reinforces the significant role of the *MAPT*-containing H1 haplotype in PD across multiple populations. Our findings highlight the genetic diversity of 17q21.31 haplotypes across different populations. It further underscores the importance of including underrepresented populations in genetic analyses. Future research should focus on understanding the structural complexity of the 17q21.31 region, utilizing long-read sequencing and multi-omics approaches, and ensuring the inclusion of diverse populations to gain a more comprehensive understanding of *MAPT*-related neurodegenerative disease risk.

## Data Availability Statement

Data used in the preparation of this article were obtained from the Global Parkinson’s Genetics Program (GP2; https://gp2.org). Specifically, we used Tier 2 data from GP2 releases 6 [DOI: https://doi.org/10.5281/zenodo.10472143)] and 7 [DOI: https://doi.org/10.5281/zenodo.10962119]. Tier 1 data can be accessed by completing a form on the Accelerating Medicines Partnership in Parkinson’s Disease (AMP®-PD) website (https://amp-pd.org/register-for-amp-pd). Tier 2 data access requires approval and a Data Use Agreement signed by your institution.

## Data Sharing

All GP2 data is hosted in collaboration with the Accelerating Medicines Partnership in Parkinson’s disease, and is available via application on the website (https://amp-pd.org/register-for-amp-pd;https://doi.org/10.5281/zenodo.7904832).

Genotyping imputation, quality control, ancestry prediction, and processing was performed using GenoTools v1.0, publicly available on GitHub (https://github.com/GP2code/GenoTools). All code generated for this article, and the identifiers for all software programs and packages used, are available on GitHub [https://github.com/GP2code/MAPThaplotype] and were given a persistent identifier via Zenodo [DOI: 10.5281/zenodo.15933056].

## Acknowledgements

This work was carried out with the support and guidance of the ‘GP2 Trainee Network’ which is part of the Global Parkinson’s Genetics Program and funded by the Aligning Science Across Parkinson’s (ASAP) initiative. Data used in the preparation of this article were obtained from Global Parkinson’s Genetics Program (GP2). For a complete list of GP2 members, see https://gp2.org. Data used in the preparation of this article were obtained from the Accelerating Medicines Partnership® (AMP®) Parkinson’s Disease (AMP® PD) Knowledge Platform. For up-to-date information on the study, visit https://www.amp-pd.org. ACCELERATING MEDICINES PARTNERSHIP and AMP are registered service marks of the US Department of Health and Human Services.

## Funding

This research was supported in part by the Intramural Research Program of the NIH, National Institute on Aging (NIA), National Institutes of Health, Department of Health and Human Services; project number ZIA AG000535 and ZIA AG000949, as well as the National Institute of Neurological Disorders and Stroke (NINDS) and the National Human Genome Research Institute (NHGRI). Data used in the preparation of this article were obtained from Global Parkinson’s Genetics Program (GP2; https://gp2.org) GP2 is funded by the Aligning Science Across Parkinson’s (ASAP) initiative and implemented by The Michael J. Fox Foundation for Parkinson’s Research. Additional funding was provided by The Michael J. Fox Foundation for Parkinson’s Research through grant MJFF-009421/17483. The AMP® PD program is a public-private partnership managed by the Foundation for the National Institutes of Health and funded by the National Institute of Neurological Disorders and Stroke (NINDS) in partnership with the Aligning Science Across Parkinson’s (ASAP) initiative; Celgene Corporation, a subsidiary of Bristol-Myers Squibb Company; GlaxoSmithKline plc (GSK); The Michael J. Fox Foundation for Parkinson’s Research; Pfizer Inc.; Sanofi US Services Inc.; and Verily Life Sciences.

## Conflict of interest

Dr Morris is employed by UCL. He reports paid consultancy from Aprinoia and AI Therapeutics; lecture fees/honoraria - Movement Disorders Society. Research Grants from Parkinson’s UK, Cure Parkinson’s Trust, PSP Association, Medical Research Council, Michael J Fox Foundation. Dr Morris is a co-applicant on a patent application related to C9orf72 - Method for diagnosing a neurodegenerative disease (PCT/GB2012/052140)

## Appendix A

Members of the Latin American Research Consortium on the Genetics of PD (LARGE-PD) and collaborators in data collection and analysis of original LARGE-PD cohort published in 2021 ^8^

Douglas P. Loesch, Andrea R. V. R. Horimoto, Karl Heilbron, Elif I. Sarihan, Miguel Inca-Martinez, Emily Mason, Mario Cornejo-Olivas, Luis Torres, Pilar Mazzetti, Carlos Cosentino, Elison Sarapura-Castro, Andrea Rivera-Valdivia, Angel C. Medina, Elena Dieguez, Victor Raggio, Andres Lescano, Vitor Tumas, Vanderci Borges, Henrique B. Ferraz, Carlos R. Rieder, Artur Schumacher-Schuh, Bruno L. Santos-Lobato, Carlos Velez-Pardo, Marlene Jimenez-Del-Rio, Francisco Lopera, Sonia Moreno, Pedro Chana-Cuevas, William Fernandez, Gonzalo Arboleda, Humberto Arboleda, Carlos E. Arboleda-Bustos, Dora Yearout, Cyrus P. Zabetian, Paul Cannon, Timothy A. Thornton, Timothy D. O’Connor, Ignacio F. Mata, Federico Micheli, Emilia Gatto, Clarisa Marchetti, Marcelo Kauffman, Alejandro Pellene, Marcela Montiel, Alejandro San Juan, Carolina Villa, Emmanuel Franchello, Delson José da Silva, Francisco Eduardo Costa Cardoso, Helio Afonso Ghizoni Teive, Artur Francisco Schumacher-Schuh, Carlos Roberto de Mello Rieder, Marcus Vinicius Della Coletta, Bruno Lopes dos Santos Lobato, Egberto Reis Barbosa, Pedro Renato de Paula Brandão, Clécio de Oliveira Godeiro Júnior, Pedro Braga Neto, Ana Lucía Zuma de Rosso, Grace Helena Letro, Maria Gabriela dos Santos Ghilardi, Pedro Chana, Patricio Olguin, Gonzalo Arboleda Bustos, Ruth Eliana Pineda Mateus, Sonia Catalina Cerquera Cleves, Jorge Luis Orozco Velez, Jaime Fornaguera, Rossy Cruz Vicioso, Edison Vasquez, Susana Peña, Andrew Sobering, Reyna Duron Martinez, Alex Medina, Daniel Martinez Ramirez, Mayela Rodriguez, Renteria Miguel, Julia Esther Rios Pinto, Ivan Fernando Cornejo Herrea, Edward Ochoa Valle, Nicanor Mori, Angel Viñuela

## SUPPLEMENTARY MATERIAL

Figures: https://docs.google.com/document/d/1ZZUoyWdQbJONGst9JrWIfevI4SYNuD5aXoCwSjJiLG0/edit

Tables: https://docs.google.com/spreadsheets/d/1GVNBp4Thu7TaQ6X4B-m_d59z3512MjOEmQyozeZlhiw/edit?gid=0#gid=0

## Notes

### Author Declarations

All GP2 data is hosted in collaboration with the Accelerating Medicines Partnership in Parkinson's disease, and is available via application on the website (https://amp-pd.org/register-for-amp-pd;https://doi.org/10.5281/zenodo.7904832). Genotyping imputation, quality control, ancestry prediction, and processing was performed using GenoTools v1.0, publicly available on GitHub (https://github.com/GP2code/GenoTools). All code generated for this article, and the identifiers for all software programs and packages used, are available on GitHub [https://github.com/GP2code/MAPThaplotype] and were given a persistent identifier via Zenodo [DOI: 10.5281/zenodo.15933056]. ®

